# NFL COVID-19 Return to Sport Injury Prevalence Comparison

**DOI:** 10.1101/2021.12.16.21267811

**Authors:** Troy Puga, Josh Schafer, Prince Agbedanu, Kevin Treffer

**Affiliations:** Kansas City University; University of Kansas; Friends University

## Abstract

**Purpose:** During the COVID 19 Pandemic, the NFL teams have been reported to have limited training sections in preparation for their games. This study compares the prevalence of injury during the 2018, 2019, and 2020 NFL seasons, with the aim to determine the potential causes of the differences in prevalence.

**Method:** Official injury reports from each team were counted during the 17-week regular season of each year (2018, 2019, and 2020). The data was analyzed using an unpaired t-test to compare the injury prevalence between each of the three seasons.

**Results:** The 2018 season produced a total of 1,561 injuries and a mean of 48.78 injuries per team. The 2019 season produced a total of 1,897 injuries and mean of 59.28 injuries per team, while the 2020 season produced a total of 2, 484 injuries and mean of 77.63 injuries per team. An unpaired t-test was performed using the data to compare the mean number of injuries per team of each of the seasons. Comparison of the 2020 season against the 2019 season showed a statistically significant difference (P=.0003). Comparison of the 2020 season to the 2018 season found a statistically significant difference (P=.0001). Comparison between the 2019 and the 2018 seasons found a statistically significant difference (P=.0314).

**Conclusion:** Although the 2019 and 2018 season showed a statistically significant difference (P=0.0314), this difference is not as astronomical when we compare the 2020 seasons vs 2019 and 2018 seasons (P=0.0003 and P=0.0001, respectively) (**Figure 2**). The astronomical increase in injury prevalence in the 2020 season over the previous years does raise the possibility that there was reduced physiological adaption to stress, due to the limited amount of training embarked on. The limited amount of training was the result of closure of practice facilities during the COVID-19 pandemic. More physiological investigation involving players must be done at the professional and amateur levels to determine if there is a lack of physiological adaptation due to limited use of practice and training facilities during the COVID-19 pandemic.

## Introduction

The National Football League (NFL) is a professional American football league composed of 32 teams. The NFL is composed of high level elite athletes who are able to train excessively and consistently at state of the art facilities, with some of the best trainers and medical professionals in the world. Even though no amount of training can completely exclude an athlete from injuries, it has been shown that a good amount of training does offer some amount of protection from the high impact forces placed on the players’ bodies due to bone remodeling in response to pressure [1]. Athletes exhibit physiological adaptions due to the demands of sport and training. Wolff’s Law states that bones will adapt to the degree of mechanical loading, such that an increase in loading will cause the architecture of the internal and external bone layers to become stronger [1]. Conversely, a decrease in loading will cause a decrease bone strength [1]. The duration, magnitude, and rate of force applied to the bone dictate the way in which the integrity of the bone is altered [1]. Exercise adaptations will lead to physiological changes in bone, tendons, ligaments, and muscles. Resistance exercise has been shown to create significant acute hormonal response, this acute response is important for tissue growth and remodeling [2]. Anabolic hormones such as testosterone and the superfamily of growth hormones have been shown to elevate fifteen to thirty minutes post-resistance exercise, providing an adequate stimulus is present [2].

**Figure 1:**
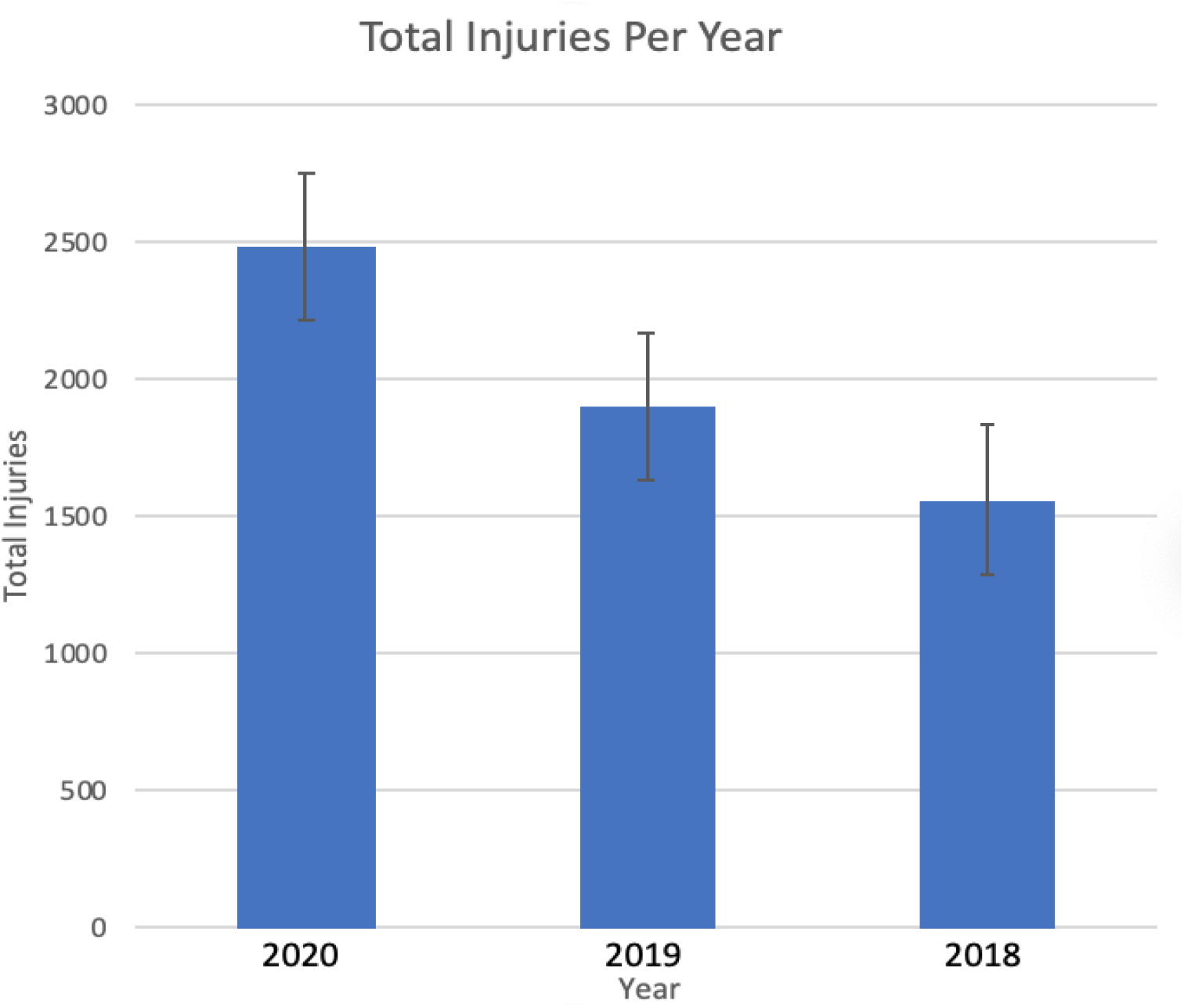
Total injuries per NFL season.

**Figure 2:**
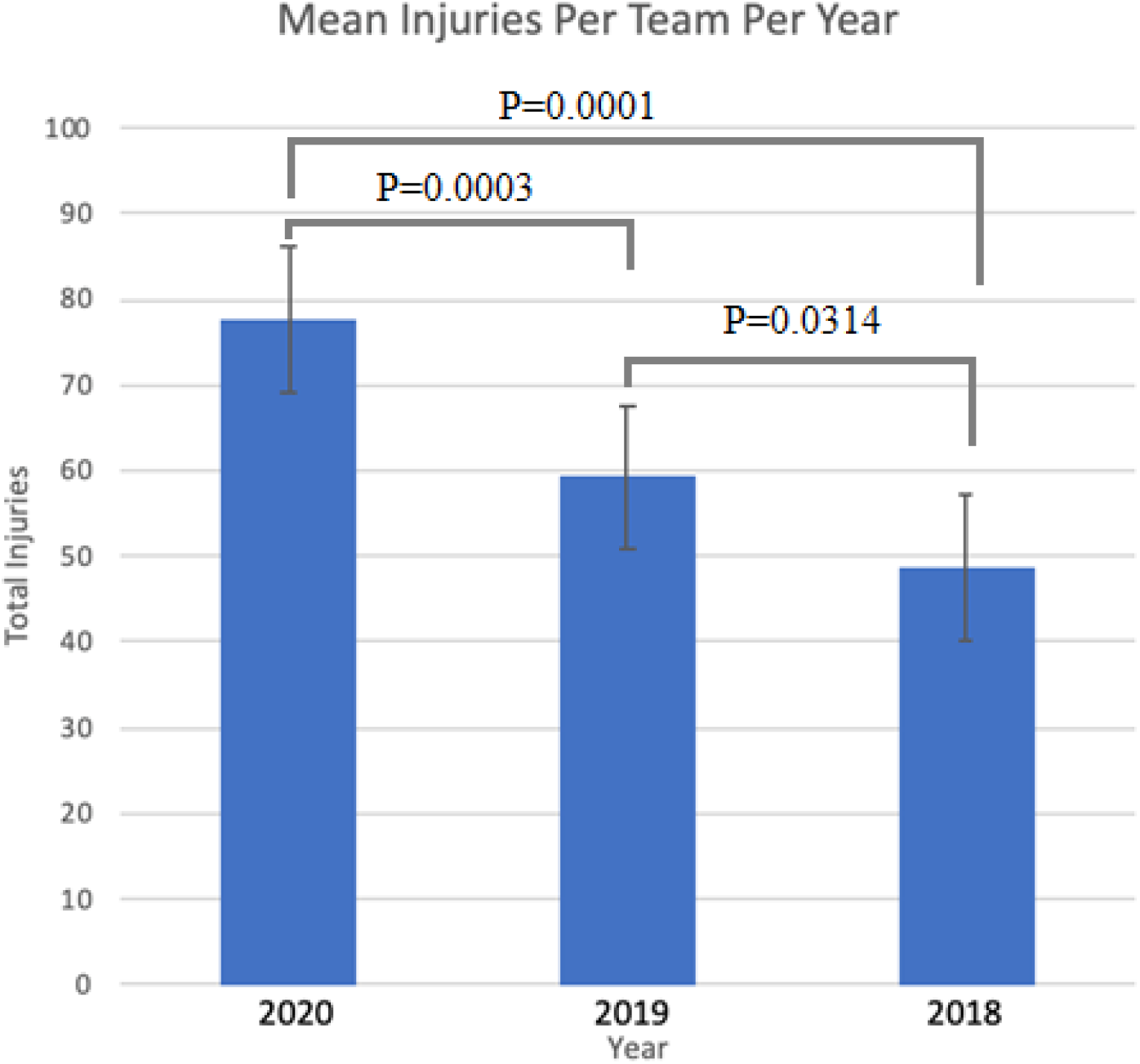
Mean injuries per team for each NFL season.

The belief is that the body will adapt to stresses, or lack of stresses placed on them, inducing pathological changes such as hyperplasia, hypertrophy, or atrophy. Increases in muscle size and strength, changes in body composition, neuroendocrine function and cardiovascular responses have been observed following resistance training [3]. In this study we aim to determine if there is a change in injury prevalence during the COVID-19 season (2020). We want to determine if a lack of sufficient training (due to restrictions on facility usage) correlates with a change in injury rates.

## Methods

The number of injuries for each team was tallied during the 17-week NFL regular season using the weekly medical data injury reports that are published publicly by the respective team. If an official team report was not available through the team, a deferral was made to the injury report on the official NFL website. Athletes on the injury reports for the same injury for consecutive weeks were only counted once, however, athletes were counted if they presented with a new injury to a different anatomical region. Illnesses, COVID-19, holidays, and non-medical days off were not included in the total tally. The total tallies per team were compared over the previous season and statistically analyzed using an unpaired t-test to compare each of the seasons to each other.

## Data analysis

A data analysis was conducted by comparing the three seasons. An unpaired t-test was selected using the data set to compare the mean number of injuries per team per season to each of the three seasons.

## Results

After the tallying of the injuries, the 2020 season produced a total 2,484 injuries with a mean of 77.625 injuries per team. The 2019 season produced a total of 1,897 injuries with a mean of 59.281 injuries per team. The 2018 season produced 1,561 injuries with a mean of 48.78 injuries per team. An unpaired t-test was performed using the data to compare the mean number of injuries per team of each of the seasons. Comparison of the 2020 season against the 2019 season showed a statistically significant difference (P=.0003). Comparison of the 2020 season to the 2018 season also showed a statistically significant difference (P=.0001). Comparison between the 2019 and the 2018 seasons showed a statistically significant difference (P=.0314) as well.

## Discussion

There could be several reasons for the difference in injury prevalence. It is believed that a decrease in physiological adaptation, due to reduced training, may contribute to the difference in injury prevalence. This could be impactful to other athletic programs that do not have the level of care provided to a player in the NFL. This information can be useful for understanding the proper protocols for sport preparation. We admit that there may also be limitations to the study in that it may not have accounted for other injuries such as preseason, injured reserve, and unreported injuries. The difference in training time must also be determined between the seasons. There must be more work done at both the professional and amateur levels to determine the level that physiological adaption plays in injury rates. Potential follow-up studies could include examining the prevalence of injuries for individual teams with COVID-19 outbreaks and examining how geographic COVID-19 hotspots impacted injuries for the teams.

## Data Availability

All data produced in the present study are available online at the official NFL website, official NFL team websites or media accounts.

https://www.nfl.com/injuries/

## Selected Resources

1. Rowe P, Koller A, Sharma S. Physiology, Bone Remodeling. [Updated 2021 Feb 9]. In: StatPearls [Internet]. Treasure Island (FL): StatPearls Publishing; 2021 Jan-. Available from: https://www.ncbi.nlm.nih.gov/books/NBK499863/

2. Kraemer, W.J., Ratamess, N.A. Hormonal Responses and Adaptations to Resistance Exercise and Training. Sports Med 35, 339–361 (2005). https://doi.org/10.2165/00007256-200535040-00004

3. Kraemer, W.J., Deschenes, M.R. & Fleck, S.J. Physiological Adaptations to Resistance Exercise. Sports Medicine 6, 246–256 (1988). https://doi.org/10.2165/00007256-198806040-00006

